# Effect of RBD mutations in spike glycoprotein of SARS-CoV-2 on neutralizing IgG affinity

**DOI:** 10.1101/2021.01.28.21250577

**Authors:** Takuma Hayashi, Nobuo Yaegashi, Ikuo Konishi

## Abstract

**Background:** Certain mutant strains of SARS-CoV-2 are known to spread widely among humans, including the receptor binding domain (RBD) mutant, Y453F, from farmed minks, and the RBD mutant, N501Y, a mutation common to three major SARS-CoV-2 subspecies (B.1.1.7, B.1.351, and B.1.1.248).

**Methods:** We investigated the characteristics of the RBD mutants, Y453F and N501Y, using three-dimensional structural analysis. We also investigated the effect of Y453F and N501Y on neutralizing antibodies in serum derived from COVID-19-positive patients.

**Results:** Our results suggest that SARS-CoV-2 subspecies with the RBD mutations Y453F or N501Y partially escaped detection by 4 neutralizing monoclonal antibodies and 21 neutralizing antibodies in serums derived from COVID-19-positive patients.

**Conclusions:** Infection with SARS-CoV-2 subspecies that cause serious symptoms in humans may spread globally.

## 1. Introduction

Mutations in the SARS-CoV-2 virus can jeopardize the efficacy of potential vaccines and therapeutics against COVID-19. Mustelidae animals (e.g., minks and ferrets) can be infected with SARS-CoV-2 relatively easily compared with other mammals [1]; however, it has not yet been elucidated why SARS-CoV-2 is extremely contagious among these animals. Nonetheless, it is clear that when several farmed minks kept in a high-density environments are infected with SARS-CoV-2, the virus proliferates in large numbers. Consequently, humans and minks may be at high risk of SARS-CoV-2 infection.

Natural selection (adaptation) in the coronavirus can occur during virus amplification *in vivo* in farmed minks [2]. Natural selection in such viruses is observed by the introduction of mutations in SARS-CoV-2 that are not observed during the growth process in humans [2,3]. Infections with the mutant strain Y453F (from farmed minks) or N501Y (a mutation common to three major subspecies of SARS-CoV-2: B.1.1.7, B.1.351, and B.1.1.248) are known to widely spread among humans [4,5,6] (Supplementary Figure 1).

In this study, we investigate the virological characteristics of these two receptor binding domain (RBD) mutants using three-dimensional protein structural analysis [7,8]. We also investigate the affinity of IgG for the conventional RBD and the mutant RBDs, Y453F and N501Y, in serum obtained from 41 COVID-19-positive patients and 20 COVID-19-negative patients. The findings indicate that SARS-CoV-2 infection with Y453F or N501Y mutations in the spike glycoprotein may escape the antiviral effect of neutralizing antibodies or COVID-19 vaccination.

## 2. Materials and Methods

### 2.1. Analysis of the three-dimensional structures of the binding sites of mink and human angiotensin-converting enzyme 2 (ACE2)

Some subspecies of SARS-CoV-2 have the amino acid mutations, Y453F or N501Y, in the sequence that encodes the spike glycoprotein [4,7,8]. These SARS-CoV-2 mutations have been detected in approximately 300 viral sequences isolated from European and Dutch populations, as well as in minks. We used the data on the three-dimensional structure of the RBD of the spike glycoprotein of SARS-CoV-2 (Protein Data Bank (PDB) ENTITY SEQ 6VW1_1) [7] and the data (PDB: 6XC2, 6XC4, 7JMP, 7JMO, 6XKQ, 6XKP, and 6XGD; Supplementary Figure 5) for the three-dimensional structure of six neutralizing antibodies (CC12.1, CC12.3, COVA2-39, COVA2-04, CV07-250, CV07-270, and REGN-COV2) (Supplementary Figure 5) that bind to the spike glycoprotein of SARS-CoV-2 [8].

Using the Spanner program, we predicted the three-dimensional structure of the SARS-CoV-2 spike glycoprotein Y453F mutant based on PDB data (ENTITY SEQ 6VW1_1). We investigated the binding of the spike glycoprotein mutant Y453F to human angiotensin-converting enzyme 2 (ACE2) and determined the affinity of the spike glycoprotein mutants, Y453F and N501Y, to six neutralizing monoclonal antibodies using the MOE program (three-dimensional protein structure modeling, protein docking analysis: MOLSIS Inc., Tokyo, Japan) and Cn3D Macromolecular Structure Viewer. Protein contact residues and buried surface areas were analyzed using the LigPlot+ program (v.1.4.5) (https://www.ebi.ac.uk/thornton-srv/software/LigPlus/). Protein buried surface areas were analyzed using the PDBePISA tool (http://pdbe.org/pisa/) and MOE project DB (MOLSIS Inc. Tokyo Japan). The modeling and docking of mink ACE2 and RBD in the spike glycoprotein of SARS-CoV-2 was analyzed by MOE project DB with previously posted ID PDB and protein ID (MOLSIS Inc. Tokyo Japan). The binding affinity between mink ACE2 and the SARS-CoV-2 spike glycoprotein RBD was analyzed using MOE project DB (MOLSIS Inc.).

### 2.2. Adsorption of conventional RBD, Y453F and N501Y recombinant proteins to the solid phase surface of the ELISA plate

HeLa cells were transfected with either pcMV3-2019-nCov-RBD-Flag tag expression vector (2 μg), pcMV3-2019-nCov-RBD Y453F-Flag tag expression vector (2 μg), or pcMV3-2019-nCov-RBD N501Y-Flag tag expression vector (2 μg) (Sino Biological Inc. Beijing, China). The cells transfected with 2019-nCov-RBD, 2019-nCov-RBD Y453F, or 2019-nCov-RBD N501Y expression vectors were incubated for 48 h prior to harvesting. The conventional RBD, Y453F, and N501Y recombinant proteins were purified using a His-tag column following the standard procedure. Purified conventional RBD, Y453F, or N501Y recombinant proteins were adsorbed on the solid phase surface of the ELISA plate (SMILON ELISA plate MS-3508F, Tokyo Japan).

### 2.3. Quantitative measurement of SARS-CoV-2 neutralizing antibodies

The SeroFlash™ SARS-CoV-2 Neutralizing Antibody Assay Fast Kit (EpiGentek Group Inc., NY) contains all the reagents necessary for quantitatively measuring the level of SARS-CoV-2 neutralizing antibodies. In this assay, conventional RBD, Y453F, or N501Y spike proteins were stably pre-coated onto microplate wells. His-tagged ACE2 binds to the coated spike protein in the presence or absence of neutralizing antibodies in the sample. The amount of the bound ACE2, which is proportional to ACE2 inhibition intensity, is then recognized by the neutralizing detection complex, which contains anti-His antibodies and is measured through an ELISA-like reaction, where the absorbance is read by a microplate spectrophotometer at a wavelength of 450 nm. The neutralizing antibody level is inversely proportional to the optical density intensity measured, i.e., the higher the level of neutralizing antibody, the lower the OD intensity. Quantitative measurements of SARS-CoV-2 neutralizing antibody levels were performed according to the manufacturer’s procedure.

### 2.4. Qualitative measurement of SARS-CoV-2 neutralizing antibodies

Serum samples (25 µL) from 20 COVID-19-negative patients and 41 COVID-19-positive patients with varying immunoglobulin M (IgM) and immunoglobulin G (IgG) antibody levels were used for this assay. COVID-19 status was confirmed by RT-PCR, antigen, and/or antibody serology tests. All serum samples were provided by RayBiotech (RayBiotech Life, GA). The details of the materials and methods used are described in the supplementary materials. A neutralizing inhibition score of ≥ 20% was used to indicate a positive result and detection of SARS-CoV-2 neutralizing antibody; a score under 20% was used to indicate a negative result and no detectable SARS-CoV-2 neutralizing antibody.

## 3. Results

### 3.1. Structural analysis

The results from the Spanner analysis revealed that the RBD mutants Y453F and N501Y did not affect the three-dimensional structure of conventional SARS-CoV-2 spike glycoproteins (Supplementary Figure 2). The results clarified that the binding property of the Y453F mutant spike glycoprotein to human ACE2 was slightly weaker than that of the conventional SARS-CoV-2 spike glycoprotein (Figure 1A, Table 1). Conversely, the binding property of the N501Y mutant spike glycoprotein to human ACE2 was stronger than that of the conventional RBD (Figure 1B, Table 1). The slightly weaker affinity observed in Y453F was due to the replacement of Tyr at position 453 by Phe, which was unable to form a hydrogen bond with His at position 34 in human ACE2 (Figure 1A). The strong affinity observed in the N501Y mutant was due to the replacement of Asn at position 501 by Tyr, which was able to form a hydrogen bond with Lys at position 353 in human ACE2, and to create a benzene ring interaction with Tyr at position 41 in human ACE2 (Figure 1B).

**Table 1.**
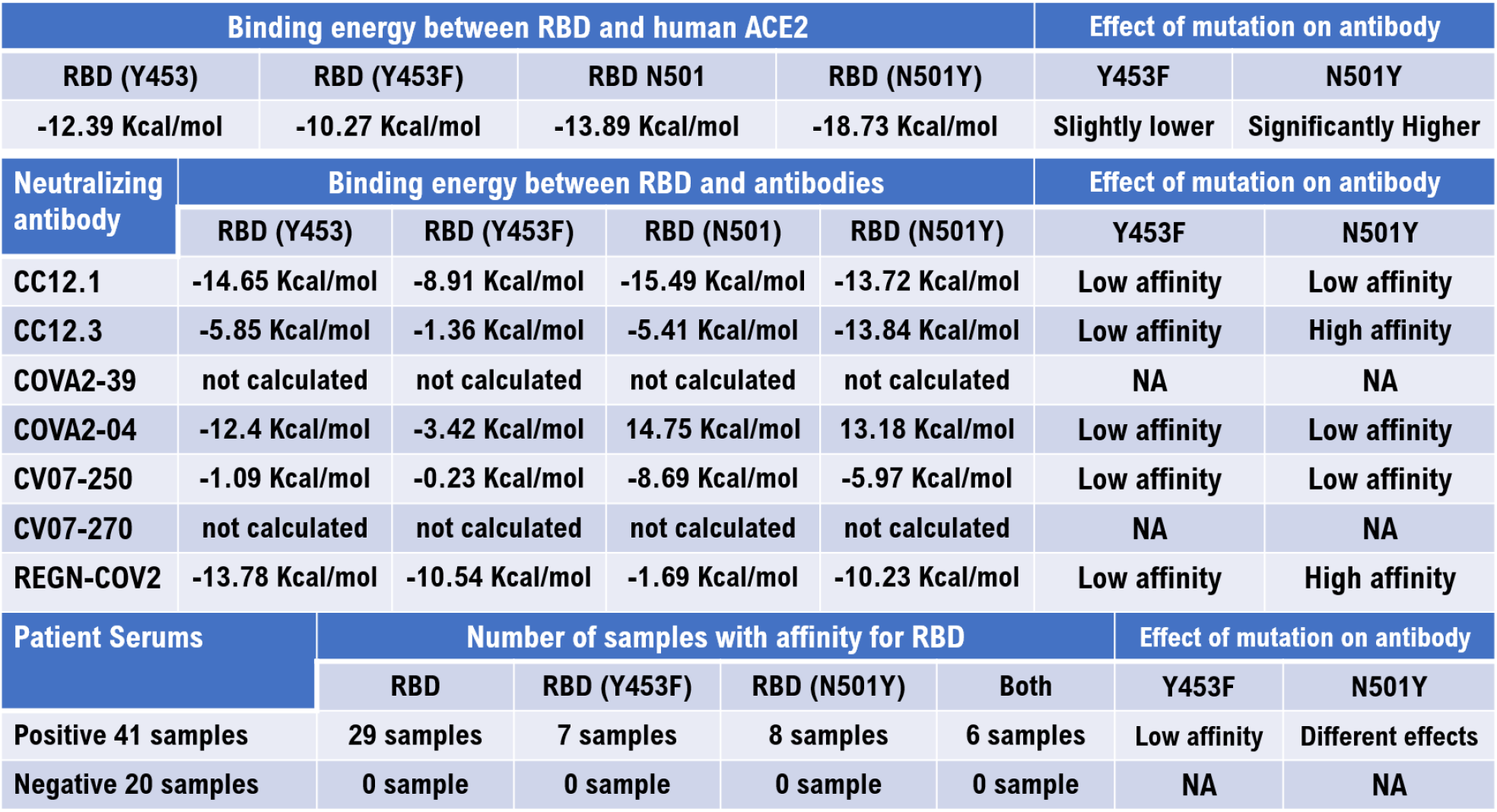
Affinity of conventional RBD, Y453F, and N501Y with neutralizing antibodies or serum from each patient. The binding of Y453F or N501Y to human ACE2 and to six neutralizing monoclonal antibodies were investigated using the MOE program and Cn3D macromolecular structure viewer. Binding energy, calculated by the MOE program, is shown in the table. Quantitative measurement of SARS-CoV-2 neutralizing antibody levels was examined using a SeroFlash™ SARS-CoV-2 Neutralizing Antibody Assay Fast Kit (EpiGentek Group Inc. NY). A strong affinity for the conventional RBD was shown in the serum IgG of 29 out of 41 COVID-19-positive patients. However, no affinity for the RBD mutant, Y453F, was shown in the serum IgG of 34 out of 41 COVID-19-positive patients. Weak affinity for both conventional RBD and N501Y was shown in the serum IgG of 7 out of 41 COVID-19-positive patients. However, no affinity for N501Y was shown in the serum IgG of 33 out of 41 COVID-19-positive patients. Various affinity for both conventional RBD and N501Y was shown in the serum IgG of 8 out of 41 COVID-19-positive. No affinity for conventional RBD, Y453F, or N501Y was shown in the serum IgG of all 20 COVID-19-negative subjects. Detailed experimental results are indicated in Supplementary Table 1.

**Figure 1:**
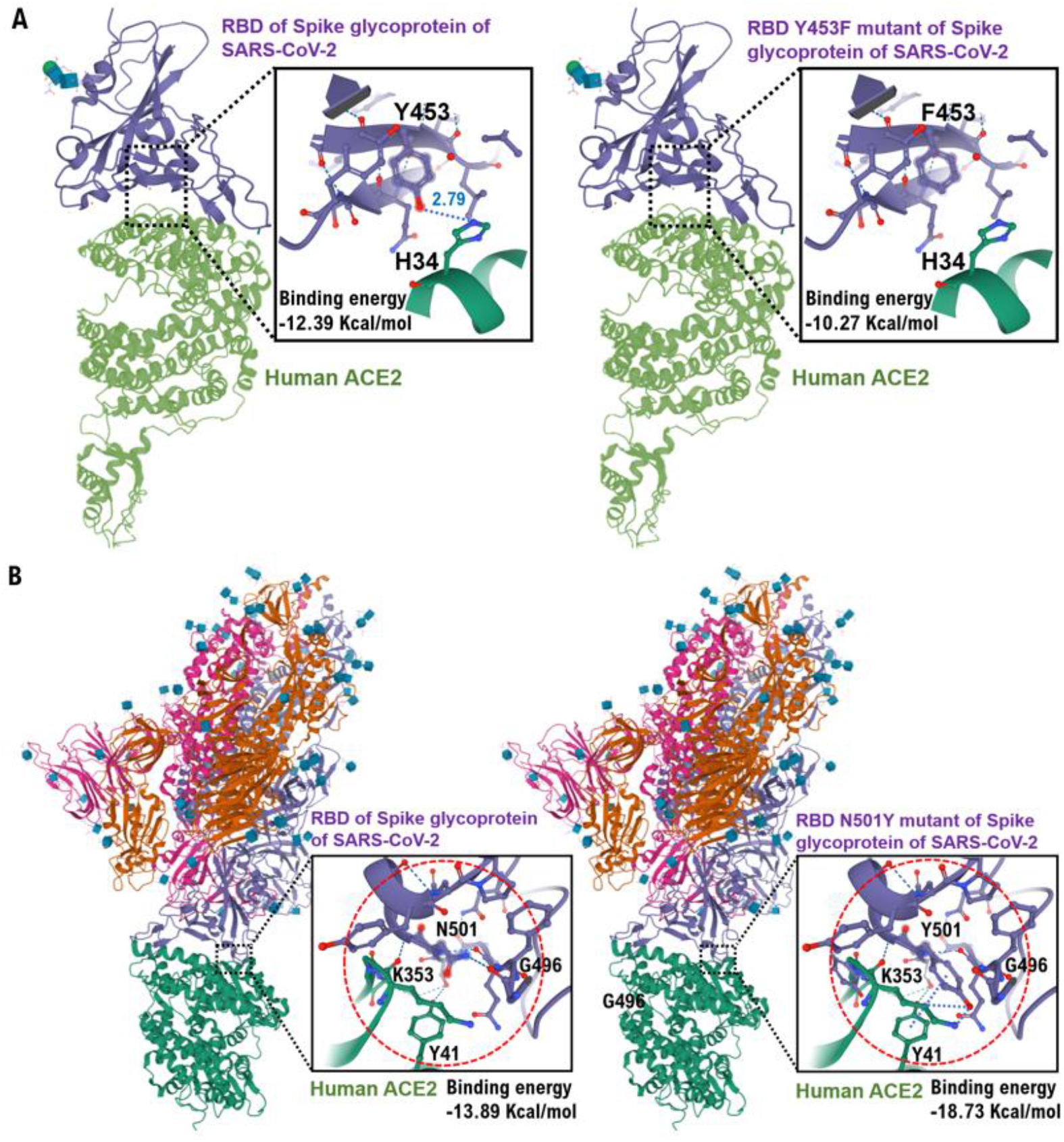
Interactions between human ACE2 and the SARS-CoV-2 spike glycoprotein receptor binding domain (RBD) mutants, Y453F and N501Y. **(A)** The interaction between ACE2 (green) and residues of the conventional RBD and the Y453F variant (purple) is shown using the three-dimensional structure model. It is speculated that the Y453 amino acid residue of the conventional RBD is hydrogen-bonded to the H34 amino acid residue of human ACE2. However, the binding between the F453 amino acid residue of the RBD mutant and the H34 amino acid residue of human ACE2 is presumed to be weak. From these results, the affinity between the spike glycoprotein RBD mutant Y453F and human ACE2 is presumed to be slightly weaker compared with the conventional RBD. **(B)** The interaction between ACE2 (green) and residues of the conventional RBD and the RBD mutant N501Y (purple) is shown using the three-dimensional structure model. It is speculated that the N501 amino acid residue of the conventional RBD is hydrogen-bonded to the Y41 and K353 amino acid residues of human ACE2. However, the binding between the Y501 amino acid residue of the RBD mutant and the Y41 and K353 amino acid residue of human ACE2 is presumed to be strong. From these results, the affinity between the RBD mutant (N501Y) of the spike glycoprotein and human ACE2 is presumed to be stronger compared with the conventional RBD. The three-dimensional structure models are shown by Cn3D macromolecular structure viewer. Detailed experimental results are indicated in the supplementary data.

The results of the structural analysis reveal that the affinity between the Y453F spike glycoprotein and four of the six examined monoclonal antibodies (CC12.1, CC12.3, COVA2-39, COVA2-04, CV07-250, CV07-270) was clearly weak compared with the conventional RBD (Figure 2A, Table 1, Supplementary Figure 3A-F, Supplementary Figure 5). The results also demonstrated that the affinity between the N501Y spike glycoprotein and four of the six examined monoclonal antibodies was clearly weak compared with the conventional SARS-CoV-2 spike glycoprotein RBD (Figure 2B, Table 1, Supplementary Figure 4A-F, Supplementary Figure 5).

**Figure 2:**
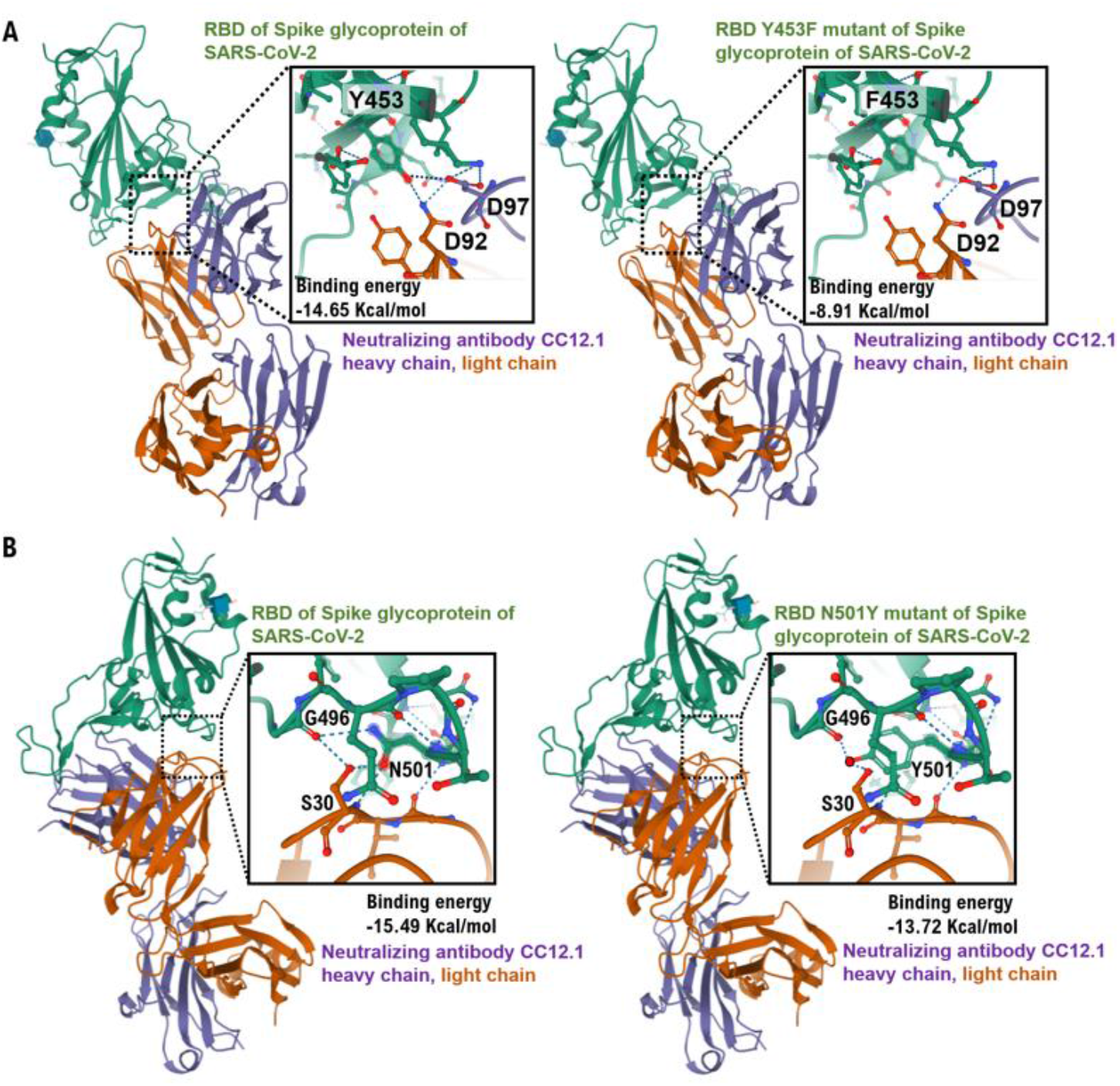
Interactions between the heavy chain of the neutralizing monoclonal antibody, CC12.2 and the SARS-CoV-2 spike glycoprotein RBD mutants, Y453F (A) and N501Y (B). **(A)** The interaction between the heavy chain (purple) and light chain (brown) of neutralizing monoclonal antibody CC12.1 and residues of the conventional RBD or Y453F (green) is shown using the three-dimensional structure model. It is speculated that the Y453 amino acid residue of the conventional RBD is hydrogen-bonded to the D92 amino acid residue of the light chain and to the D97 amino acid residue of the heavy chain of CC12.1. However, the binding between the F453 amino acid residue of the RBD mutant and the D92 and D97 amino acid residues of CC12.1 is presumed to be weak. From these results, the affinity between the spike glycoprotein of Y453F and CC12.1 is presumed to be lower compared with the conventional RBD. **(B)** The interaction between the heavy chain (purple) and light chain (brown) of CC12.1 and the residues of the conventional RBD or N501Y (green) is shown using the three-dimensional structure model. It is speculated that the N501 amino acid residue of the conventional RBD is hydrogen-bonded to the S30 amino acid residue of the light chain of CC12.1. However, the binding between the Y501 amino acid residue of the RBD mutant and the S30 amino acid residue of CC12.1 is presumed to be weak. From these results, the affinity between the RBD of N501Y and CC12.1 is presumed to be slightly lower compared with the conventional RBD. The three-dimensional structure models are shown by Cn3D macromolecular structure viewer. Detailed experimental results are indicated in the supplementary data.

### 3.2. Quantitative measurement of SARS-CoV-2 neutralizing antibodies

On September 28, 2020, the US pharmaceutical manufacturer, Regeneron Pharmaceuticals, announced the production of the antibody cocktail therapy, REGN-COV2, which combined two neutralizing antibodies, casirivimab and imdevimab, for the treatment and prevention of COVID-19 [9] (Figure 3A). On January 11, 2021, Regeneron Pharmaceuticals stated the likelihood of the antiviral effectiveness of REGN-COV2 on SARS-CoV-2 subspecies [10]. Therefore, we investigated the affinity of REGN-COV2 to the spike glycoproteins of Y453F and N501Y. We found that the affinity of REGN-COV2 (6XGD) to Y453F was weaker than that to the conventional RBD (Figure 3B, Table 1). However, the affinity of REGN-COV2 to N501Y was strong compared to the conventional RBD (Figure 3B). Therefore, the antiviral effect of REGN-COV2 as a neutralizing antibody may be maintained even against RBD mutant strains of SARS-CoV-2.

**Figure 3.**
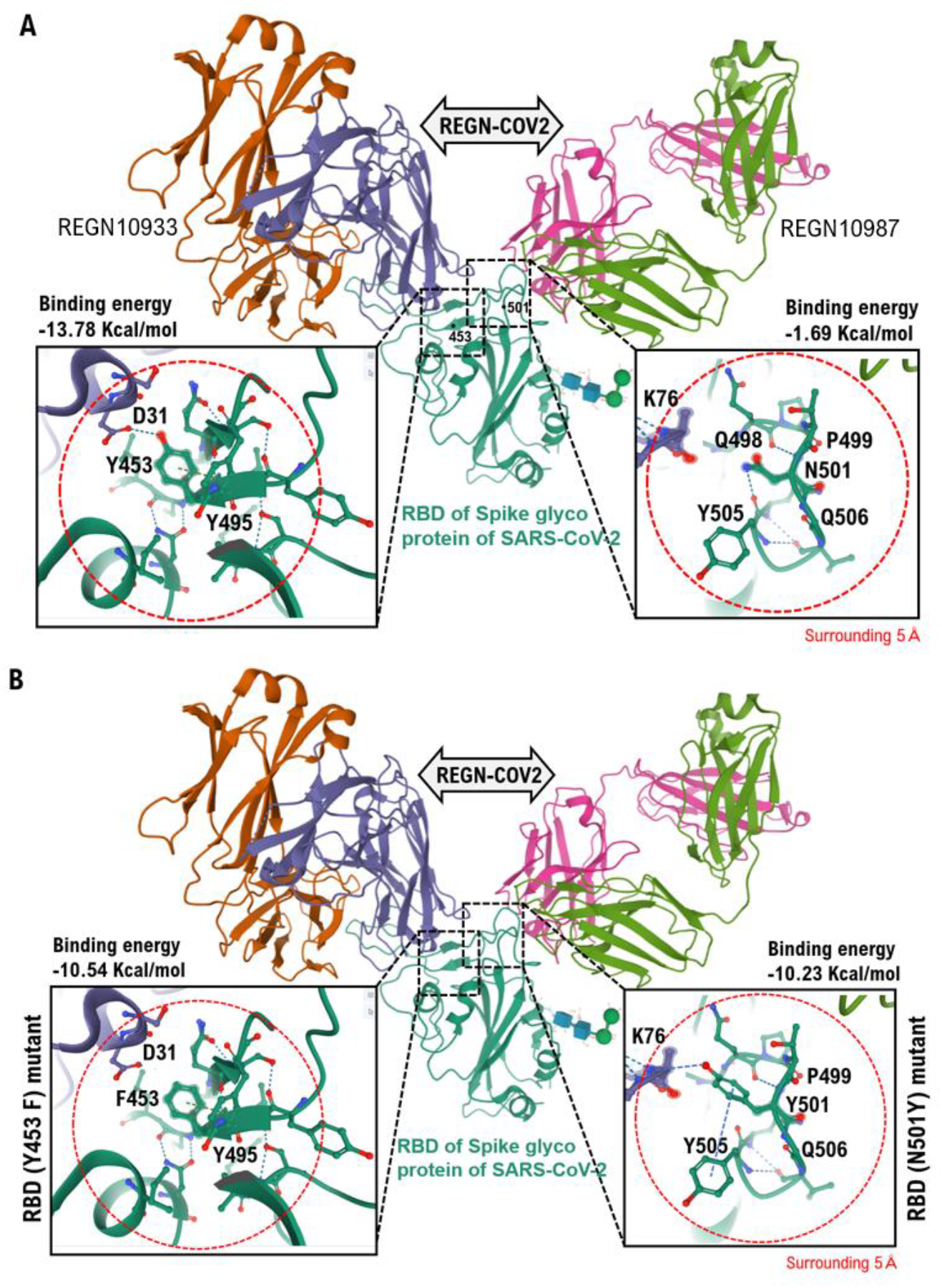
Interactions between the antibody cocktail therapy REGN-COV2 (casirivimab REGN10933 and imdevimab REGN10987; 6XGD) and Y453F and N501Y. **(A)** The interaction between the heavy chains (purple, light green) and light chains (brown, pink) of casirivimab and imdevimab and residues of the conventional RBD (green) is shown using the three-dimensional structure model. It is speculated that the Y453 amino acid residue of the conventional RBD is hydrogen-bonded to the D31 amino acid residue of the light chain of casirivimab. However, the binding between the N501 amino acid residue of the conventional RBD and the amino acid residues of imdevimab is presumed to be weak. **(B)** The interactions between the heavy chain (purple) and light chain (brown) of casirivimab and imdevimab and residues of Y543F and N501Y (green) are shown using the three-dimensional structure model. The binding between the F453 amino acid residue of Y453F and the amino acid residue of the light chain of casirivimab is presumed to be weak. However, it is speculated that the Y501 amino acid residue of N501Y is hydrogen-bonded to the K76 amino acid residue of the light chain of imdevimab. The three-dimensional structure models are shown by Cn3D macromolecular structure viewer. Detailed experimental results are indicated in the supplementary data.

### 3.3. Qualitative measurement of SARS-CoV-2 neutralizing antibodies

The results obtained by computer analysis are only theoretical results and do not show all the medical evidence in clinical practice. Therefore, we investigated the affinity of serum IgG to the conventional RBD, Y453F, and N501Y in COVID-19-positive patients and healthy subjects. A strong affinity for conventional RBD was shown for the serum IgG of 29 of the 41 COVID-19-positive patients (Table 1, Supplementary Table 1). Moderate affinity for conventional RBD, Y453F, and N501Y was shown in the serum IgG of four of the COVID-19-positive patients (Table 1, Supplementary Table 1). No affinity for conventional RBD, Y453F, or N501Y was shown in the serum IgG of the COVID-19-negative subjects (Table 1, Supplementary Table 1).

From the results, we concluded that the mutation of tyrosine at amino acid residue 453 to phenylalanine or the mutation of asparagine at amino acid residue 501 to tyrosine eliminated the inhibitory effects of the neutralizing antibody on binding between ACE2 and the RBD of the SARS-CoV-2 spike glycoprotein. It is possible that the affinity between the appropriate amino acid residues in the variable region of the antibody and the RBD of Y453F or N501Y was diminished owing to weak recognition of the monoclonal antibody to SARS-CoV-2 spike glycoproteins.

## 4. Discussion

To the best of our knowledge, data for SARS-CoV-2 mutants have not yet been published. Therefore, it is unclear whether the SARS-CoV-2 mutants detected in people working on mink farms are actually derived from farmed minks. However, in the present study, the subspecies of SARS-CoV-2 derived from farmed minks or humans was observed in a group of infected people and those that were inherited by infected individuals (Supplementary Figure 5).

Mutations in SARS-CoV-2 that have led to the generation of SARS-CoV-2 subspecies have made humans and animals susceptible to infection through easy propagation in the host, thereby making it difficult to identify the effects of therapeutic agents or vaccines for COVID-19. Moreover, the spread of SARS-CoV-2 subspecies mediated by millions of infected farmed mink is uncontrolled, raising a concern that infection by SARS-CoV-2 subspecies that cause serious symptoms in humans may spread globally.

As of January 2021, the number of people infected with SARS-CoV-2 N501F subspecies, which are believed to have occurred in the United Kingdom, South Africa, and Brazil, has increased significantly in the UK and other European countries. Recent studies have shown that the infectivity of the N501Y variant is approximately 1.4–1.7 times that of previous strains of SARS-CoV-2 [11]. In addition, N501F variants have the property of easily infecting children. Currently, the question is whether N501Y variants are resistant to the COVID-19 vaccines which has been distributed in the UK, the US, and other countries. Pfizer, who created the first-to-be-approved COVID-19 vaccine (known as BNT162b2), has demonstrated the possibility of the immediate production of mRNA that should correspond to SARS-CoV-2 mutations. On January 8, 2021, Pfizer and BioNTech reported the efficacy of the COVID-19 vaccine against the UK and South Africa SARS-CoV-2 variants based on the results of phase I clinical trials [12]. On January 11, 2021, Regeneron Pharmaceuticals announced the antiviral effectiveness of the antibody cocktail therapy, REGN-COV2, against SARS-CoV-2 subspecies. However, concerns remain regarding the effectiveness of the COVID-19 vaccine against these SARS-CoV-2 variants [3,4,13,14,15].

Mankind has struggled to develop therapies against AIDS to correspond to the speed of mutations that arise in the human immunodeficiency virus (HIV) [16]. To date, no clinical treatment has been established that directly inhibits the SARS-CoV-2 life cycle. In the fight between humans and viruses, mankind has never been able to prevent the mutation of viruses. More than 50 countries and territories around the world currently have strict restrictions on entry from the UK, South Africa, and Brazil [17]. First, people around the world should reaffirm the importance of wearing masks, hand hygiene, and social distancing.

## Data Availability

Authors shows all data referred to in the manuscript.

## Abbreviations

AIDS: acquired immunodeficiency syndrome
COVID-19: coronavirus disease-2019
HIV: human immunodeficiency virus
IgG: immunoglobulin G
IgM: immunoglobulin M
RBD: receptor binding domain
SARS-CoV-2: severe acute respiratory syndrome coronavirus-2
UK: United Kingdom

## Footnote

All authors are receiving medical ethics education. In addition, this study has been approved as a clinical medical study at each medical facility. The human serum samples used in this study were purchased from RayBiotecn life, and therefore Informed consent from the patient is not required.

## Data Sharing

Data are available on various websites and have also been made publicly available (more information can be found in the first paragraph of the Results section).

## Ethics statement

This study was reviewed and approved by the Central Ethics Review Board of the National Hospital Organization of Japan (Meguro, Tokyo, Japan). The approved number for this study is 50-201504. In order to carry out this research, the authors attended a research ethics education course (e-APRIN) conducted by Association for the Promotion of Research Integrity (APRIN; Shinjuku, Tokyo, Japan). The approved numbers of e-APRIN are AP0000151756, AP0000151757, AP0000151758, AP0000151769.

## Acknowledgments

We thank Professor Richard A. Young (Whitehead Institute for Biomedical Research, Massachusetts Institute of Technology, Cambridge, MA) for his research assistance. This study was supported in part by grants from the Japan Ministry of Education, Culture, Science and Technology (No. 24592510, No. 15K1079, and No. 19K09840); Foundation of Osaka Cancer Research; Ichiro Kanehara Foundation for the Promotion of Medical Sciences and Medical Care; Foundation for Promotion of Cancer Research; Kanzawa Medical Research Foundation; Shinshu Medical Foundation; and Takeda Foundation for Medical Science.

## Author Contributions

T.H. performed most of the experiments and coordinated the project. T.H. and N.Y. conceived the study and wrote the manuscript. N.Y. and I.K. provided with information on clinical medicine and oversaw the entire study.

## Transparency Document

The transparency document associated with this article can be found in the online version at http://.

