## supplemental materials for "Effect of RBD mutations in spike glycoprotein of SARS-CoV-2 on neutralizing IgG affinity"

**Materials and Methods**

**Analyzes the three-dimensional structure of the binding site between mink and human ACE2**

**Spanner** is a structural homology modeling pipeline that threads a query amino-acid sequence onto a template protein structure. Spanner is unique in that it handles gaps by spanning the region of interest using fragments of known structures.

To create a model, you must provide a template structure, as well as an alignment of the query sequence you wish to model onto the template sequence. Spanner will replace mismatched residues, and fill any gaps caused by insertions or deletions.

For users that are unable to create an alignment a method for building a model starting only from sequence is also available. During this process a template search is conducted and an alignment is built dynamically using FORTE before being passed through to the main part of the pipeline.

Spanner consists of several modules written in the Go programming language. For Spanner jobs which build a model only from sequence, the first step is a search of the PDB for possible templates using BLAST. These possible templates are then aligned and scored with FORTE.

The next step involves defining the start and end points of fragments corresponding to insertions or deletions. The start and end points are referred to as anchors because they must be equivalent in both the template and any candidate fragment. The margin parameter determines how far from the edge of a gap the fragment begins or ends. For example a margin of 0 would mean that the anchors begins at the very edge of a gap. This is usually not a good idea, and the default margin is set to 1.

A representative set of protein chains was prepared using CD-HIT at 100% sequence identity.[3] All continuous fragments were then extracted from this set of chains and stored in a relational database, indexed by the internal coordinates of the fragment endpoints. A separate database is prepared for each fragment length. Currently, fragments of length 8-40, including the 8 anchor residues, are stored in the database.

**Cn3D** ("see in 3D") is a helper application for your web browser that allows researcher to view 3-dimensional structures from NCBI's Entrez Structure database. Cn3D is provided for Windows and Macintosh, and can be compiled on Unix. Cn3D simultaneously displays structure, sequence, and alignment, and now has powerful annotation and alignment editing features. *(For those who prefer to view 3D structures on the web, without the need to install a separate application, iCn3D ("I see in 3D") is available as of April 2016.)*

•**Web-based Structure Viewer**iCn3D ("I see in 3D"), released in April 2016, provides interactive views of three-dimensional macromolecular structures on the web.

•There is no need to install a separate application in order to use iCn3D; you just need to use a web browser that supports WebGL.

•iCn3D also allows you to cutomize the display of a structure and generate a URL that allows you to share the link, and to incorporate iCn3D into your own web pages.

•**New Features in Cn3D 4.3.1:**View superpositions of structures that have similar molecular complexes, as identified by the newly released VAST+ (an enhanced version of the existing Vector Alignment Search Tool). The VAST+ help document provides additional details about the tool and examples of how it can be used to learn more about proteins.

•Cn3D 4.3.1 uses the MIME type: application/vnd.ncbi.cn3d.

Analyses of protein contact residues and protein buried surface areas Protein contact residues were analyzed using the **LigPlot+ program (v.1.4.5)** (https://www.ebi.ac.uk/thornton-srv/software/ LigPlus/). Protein buried surface areas were analysed using **PDBePISA tool** (http://pdbe.org/pisa/) and **MOE project DB** (MOLSIS Inc. Tokyo Japan). The modeling and Docking of the mink ACE2 protein and RBD in Spike Glycoprotein SARS-CoV-2 was analyzed by **MOE project DB** with previously posted ID PDB and protein ID (MOLSIS Inc. Tokyo Japan). The binding affinity between mink ACE2 and RDB in Spike Glycoprotein of SRARS-CoV-2 was analyzed by **MOE project DB** (MOLSIS Inc.).

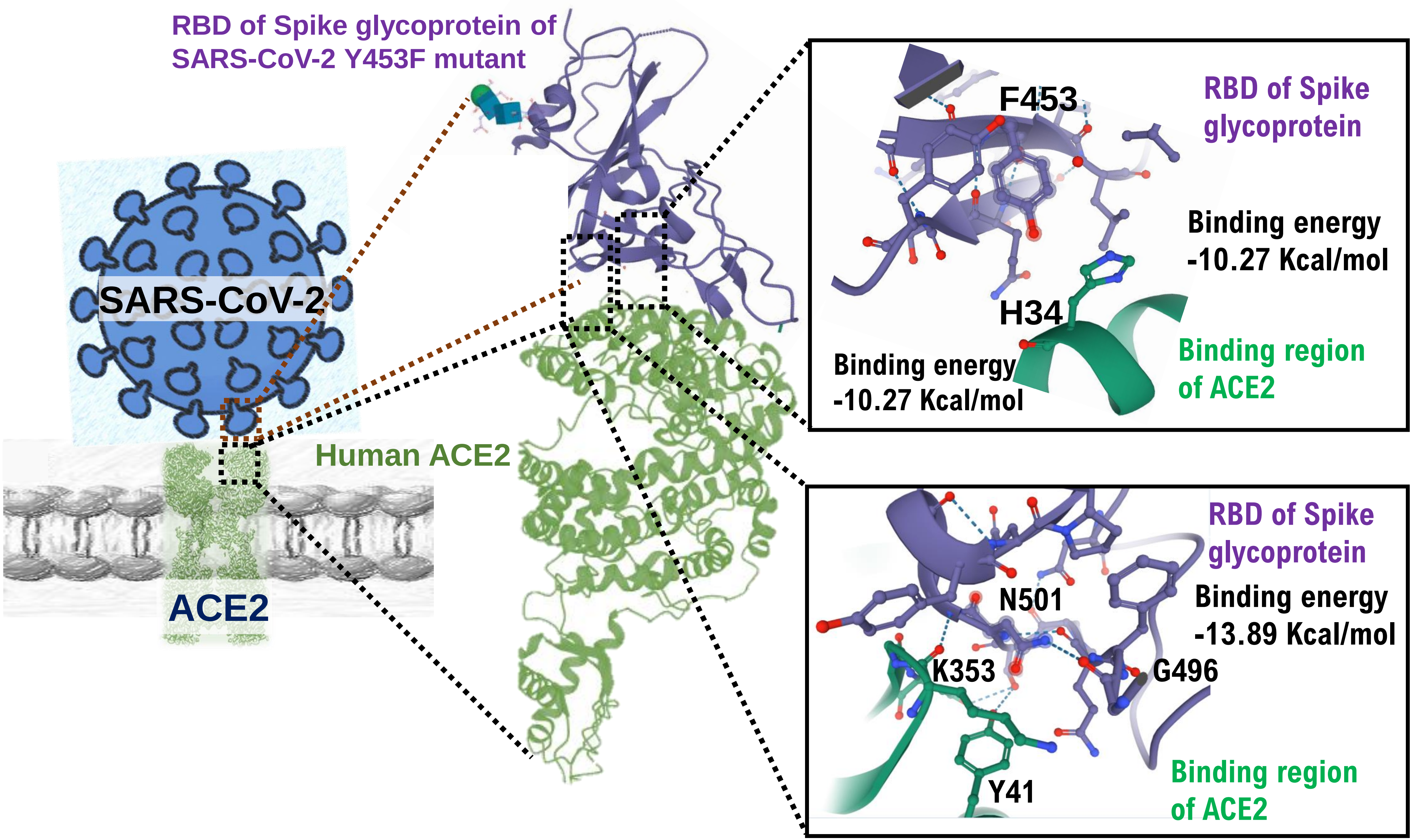
RBD of Spike glycoprotein of SARS-CoV-2 Y453F mutant
SARS-CoV-2
Human ACE2
ACE2
F453
RBD of Spike glycoprotein
Binding energy -10.27 Kcal/mol
H34
Binding energy -10.27 Kcal/mol
Binding region of ACE2
RBD of Spike glycoprotein
N501
Binding energy -13.89 Kcal/mol
K353
G496
Y41
Binding region of ACE2

Hayashi *et al*. S.Figure 2

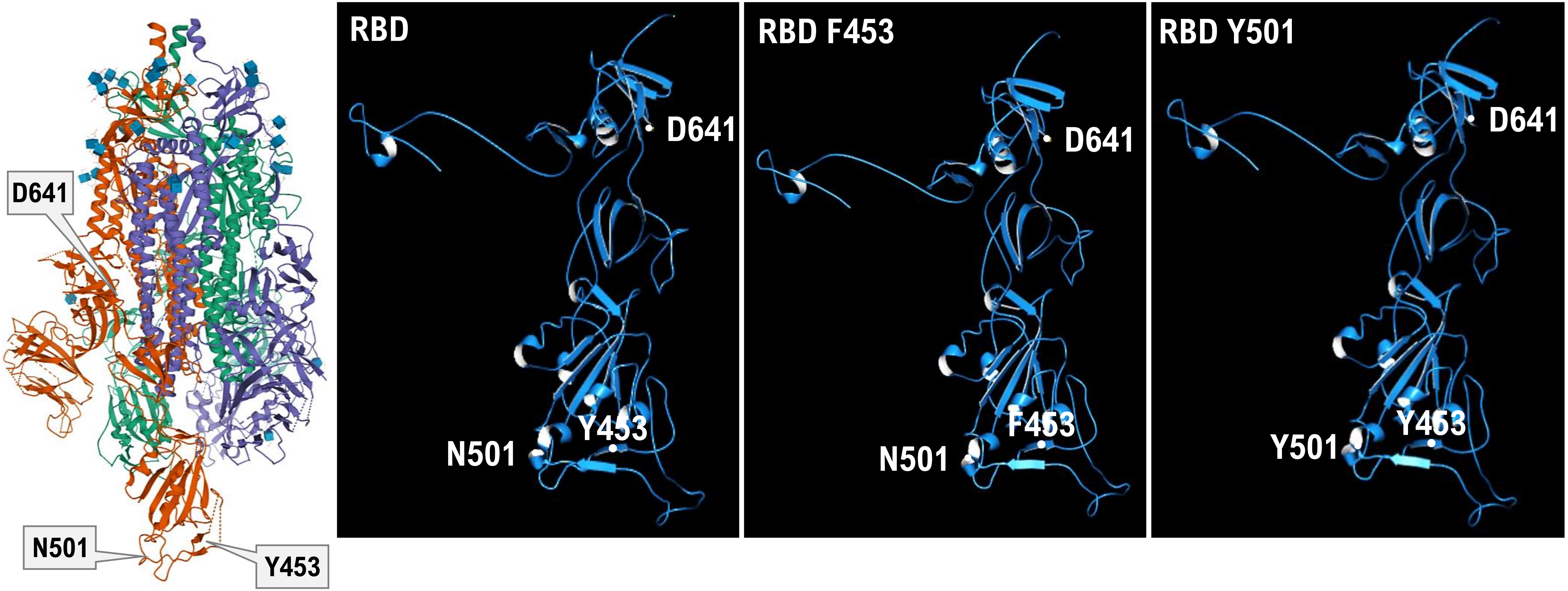


https://www.rcsb.org/3d-view/7AD1

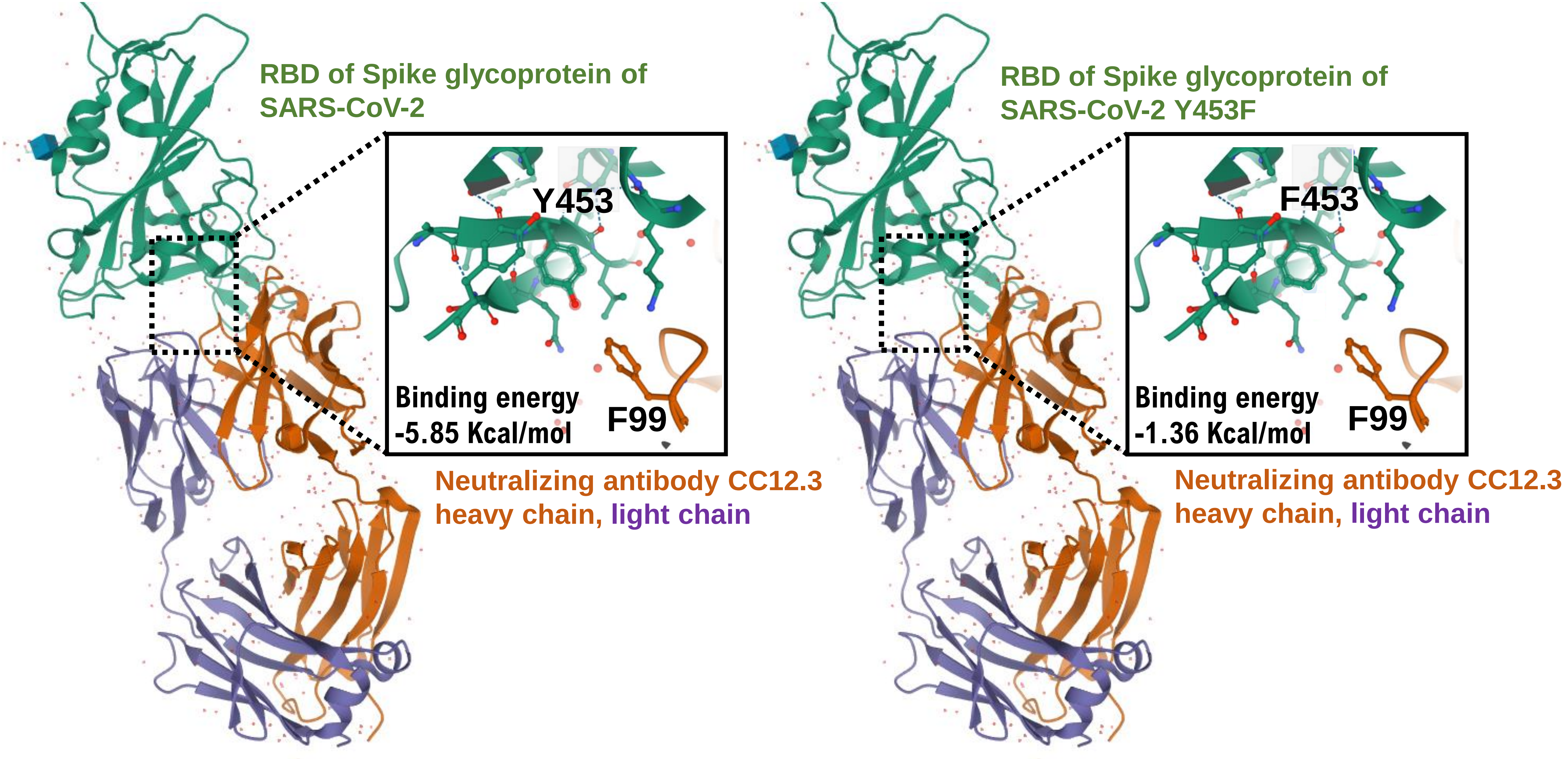


**Affinity between RBD Y453F mutant of Spike glycoprotein of SARS-CoV-2 and neutralizing antibody (CC12.3) analyzed using MOE project DB and Cn3D 4.3.1.**

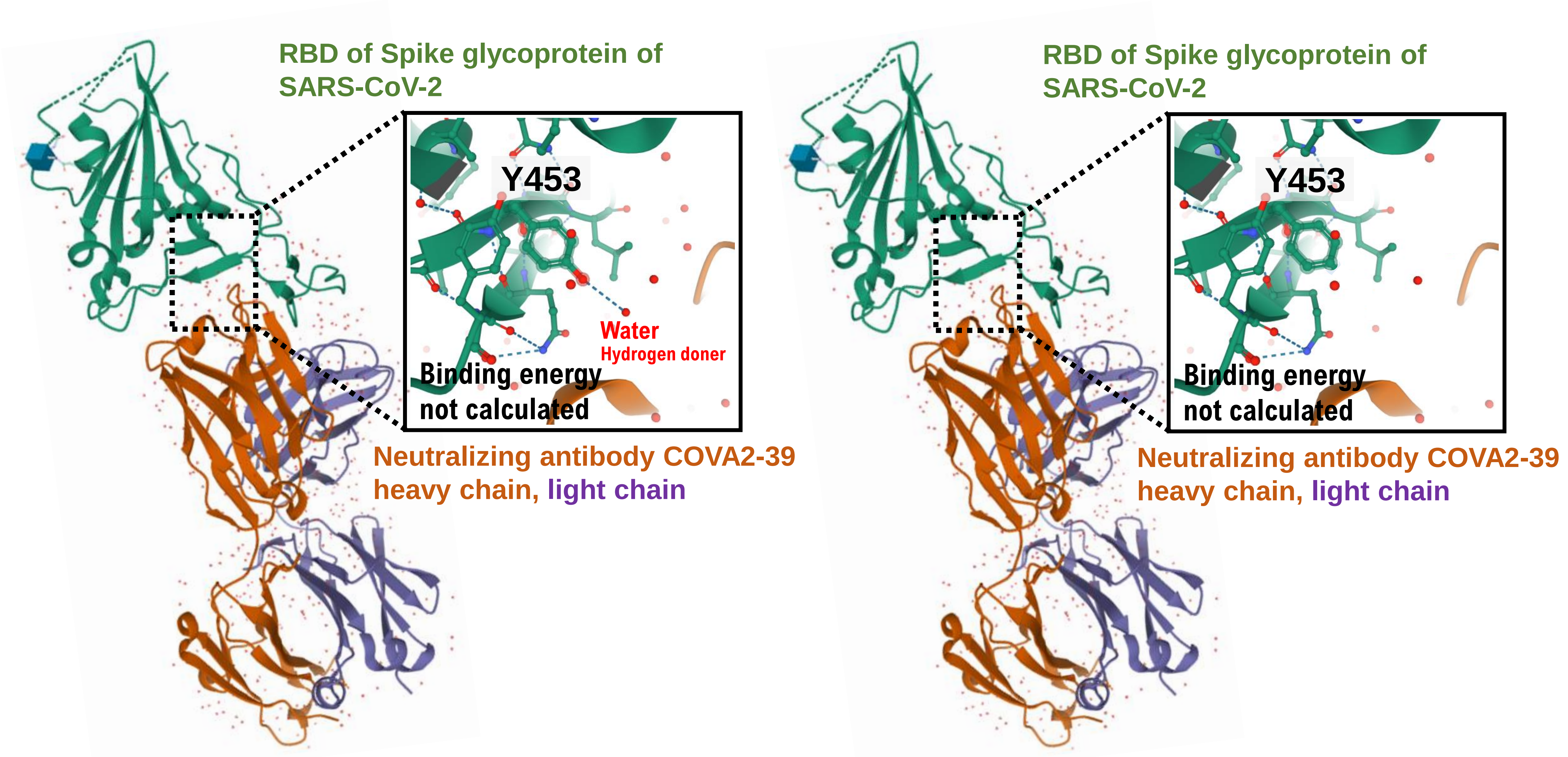


**Affinity between RBD Y453F mutant of Spike glycoprotein of SARS-CoV-2 and neutralizing antibody (COVA2-39) analyzed using MOE project DB and Cn3D 4.3.1.**

Hayashi *et al*. S.Figure 3C

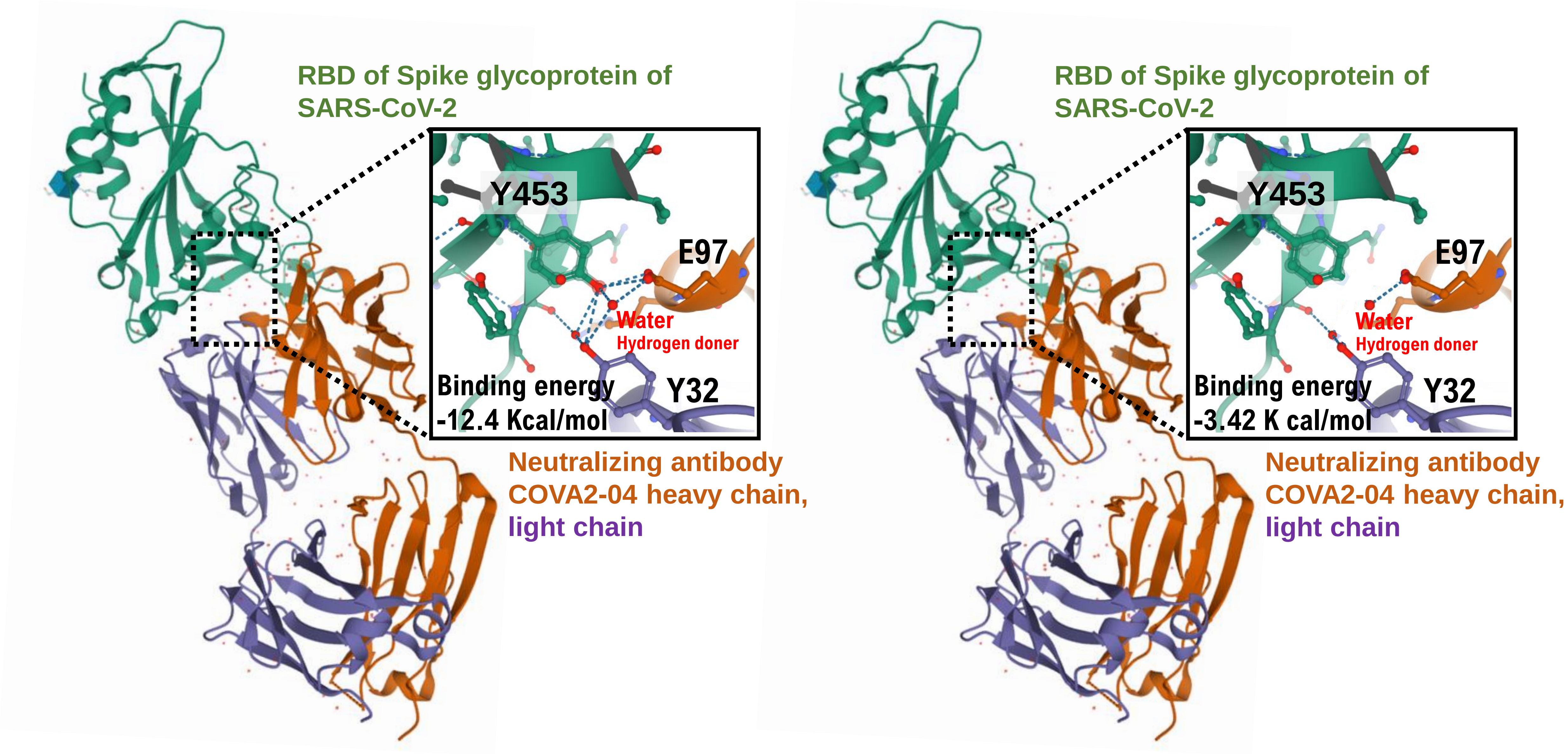


**Affinity between RBD Y453F mutant of Spike glycoprotein of SARS-CoV-2 and neutralizing antibody (COVA2-04) analyzed using MOE project DB and Cn3D 4.3.1.**

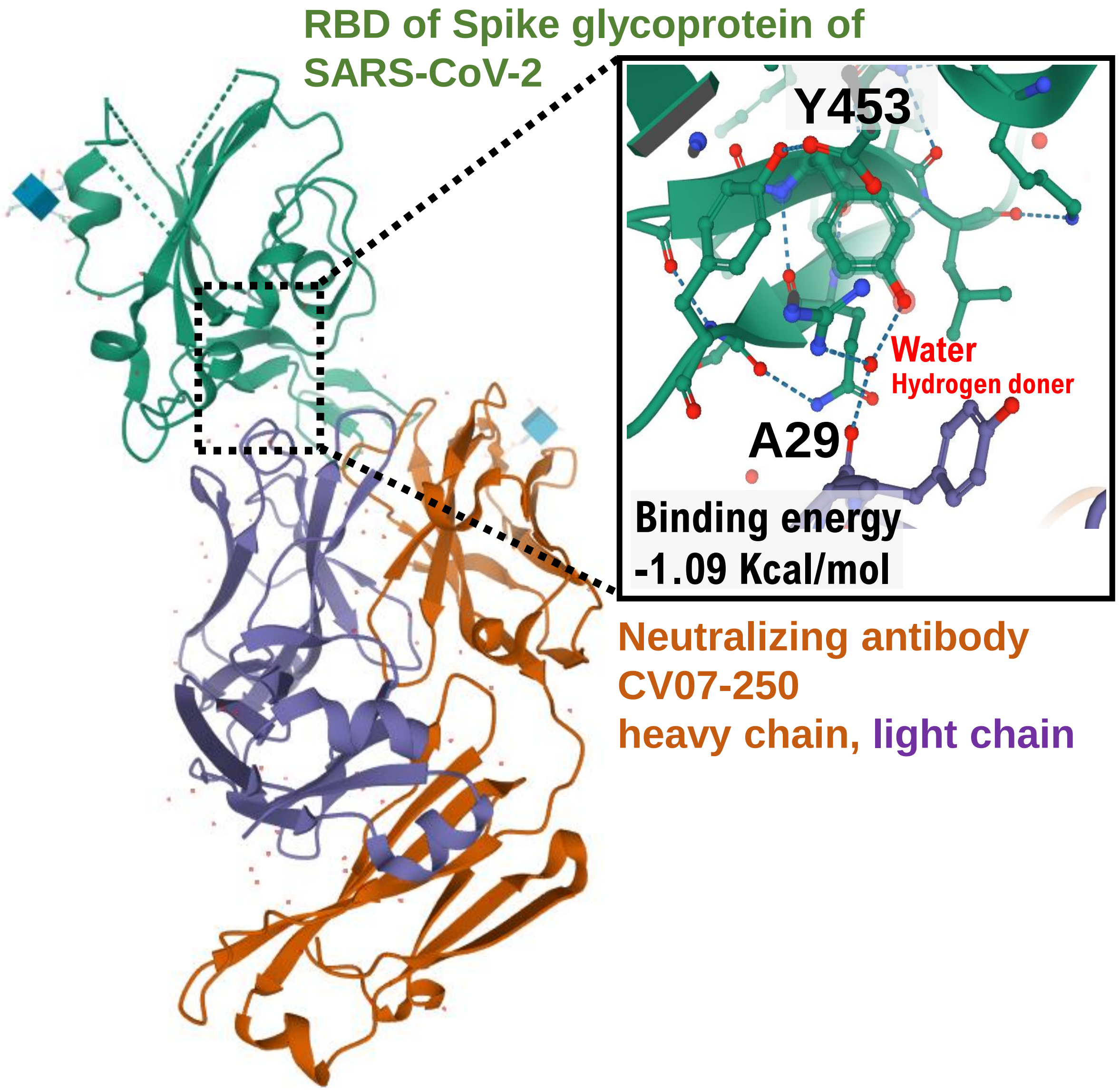


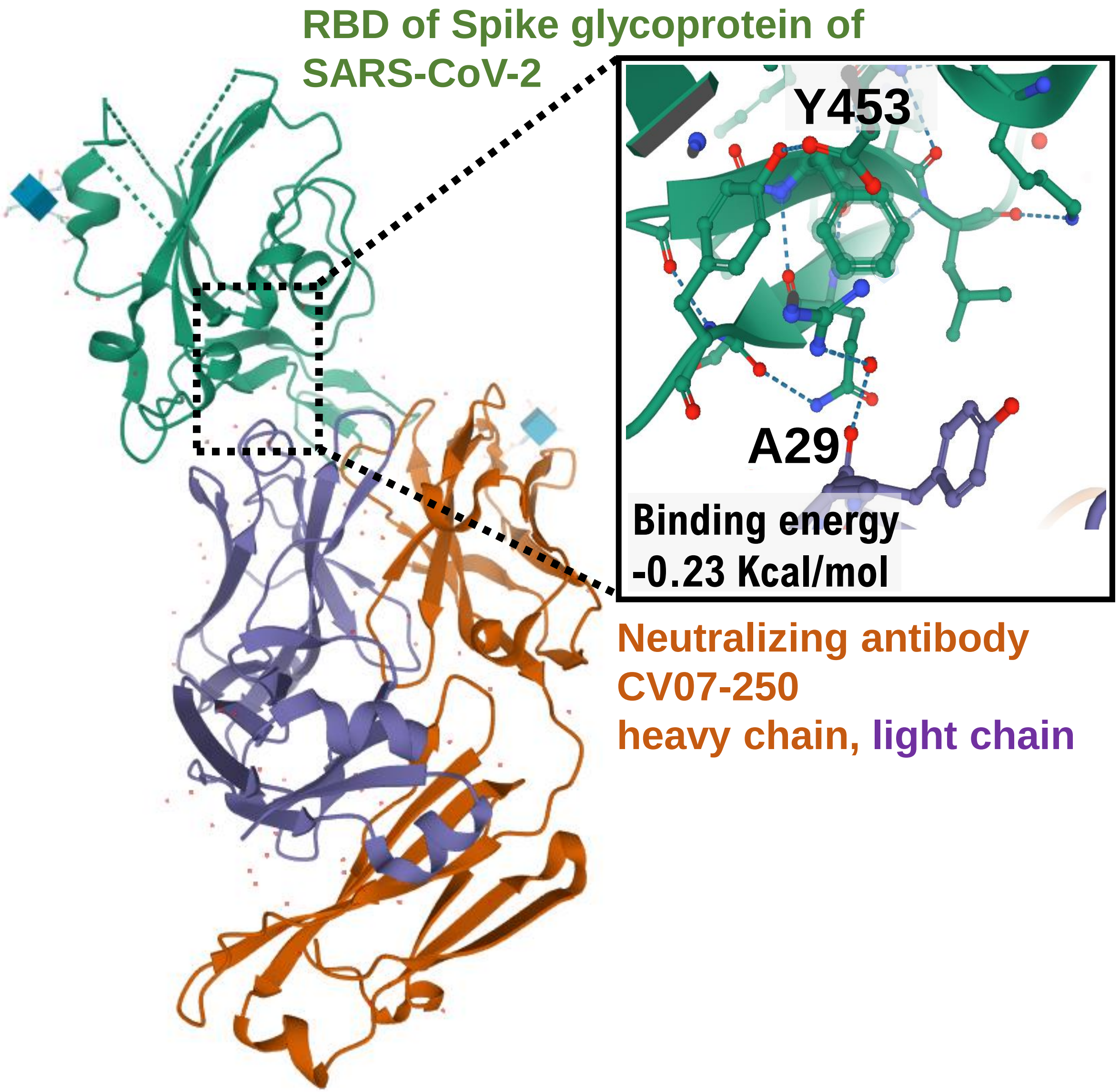


**Affinity between RBD Y453F mutant of Spike glycoprotein of SARS-CoV-2 and neutralizing antibody (CV07-250) analyzed using MOE project DB and Cn3D 4.3.1.**

Hayashi *et al*. S.Figure 3E

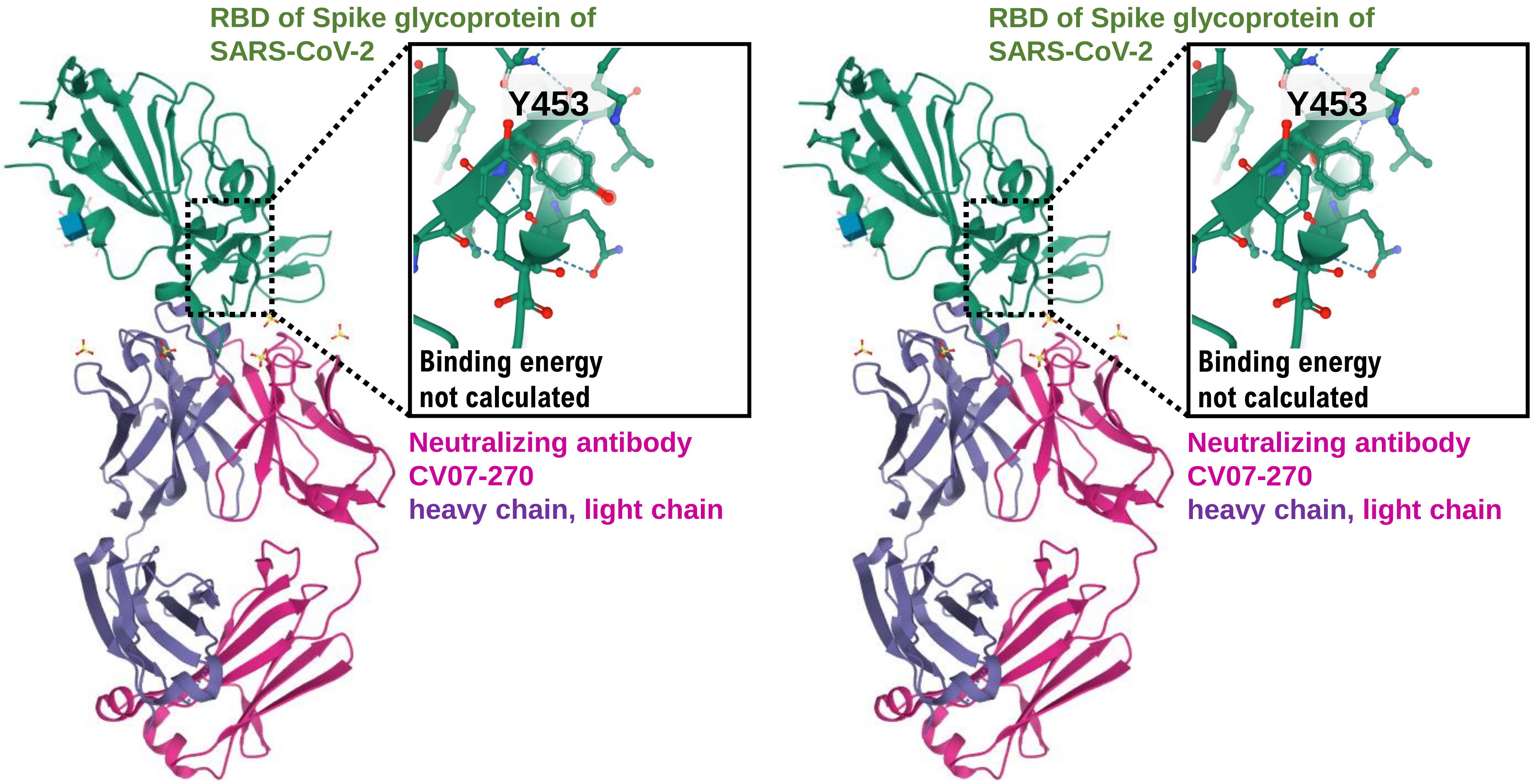


**Affinity between RBD Y453F mutant of Spike glycoprotein of SARS-CoV-2 and neutralizing antibody (CV07-270) analyzed using MOE project DB and Cn3D 4.3.1.**

Hayashi *et al*. S.Figure 3F

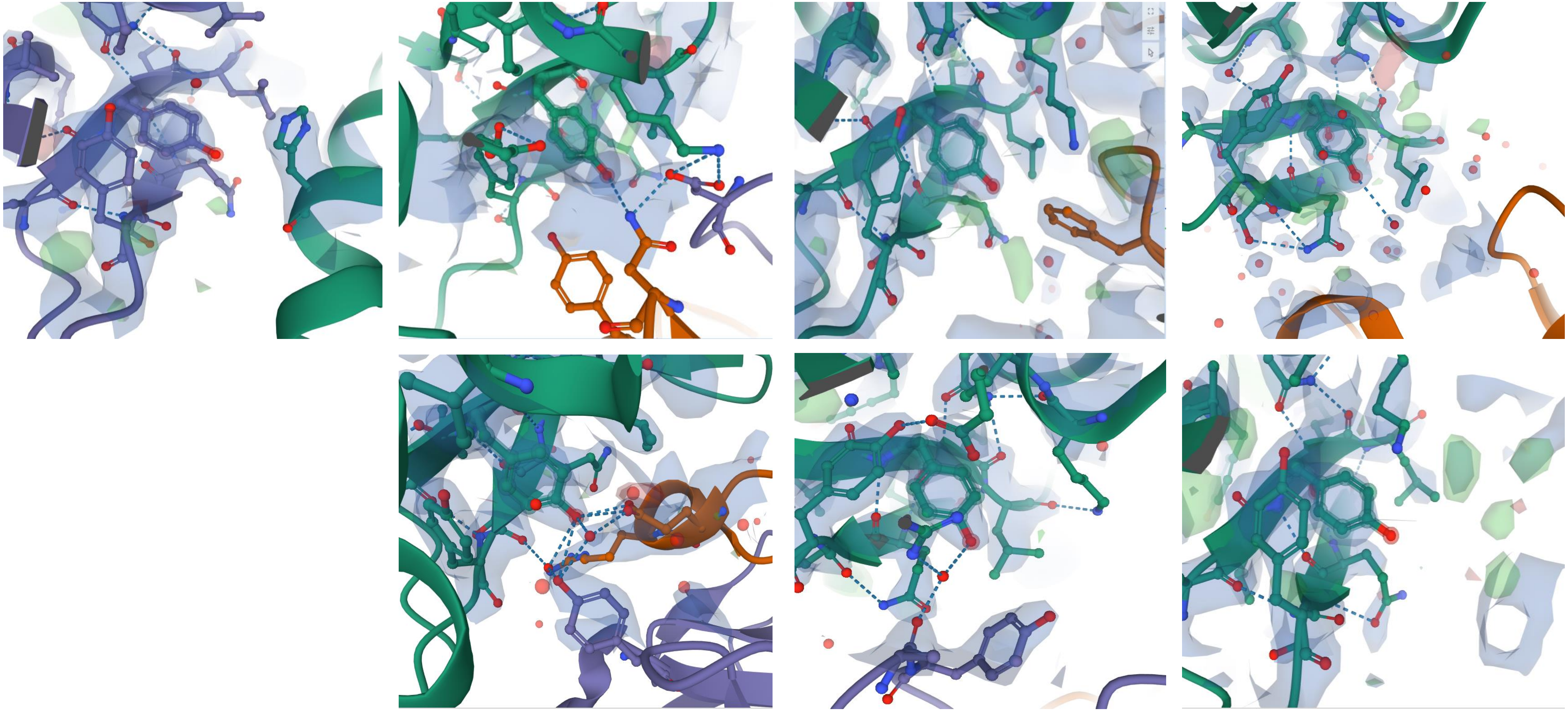

**Electron density map.** 2*F*o−*F*c electron density maps contoured at 1.5*σ* at the binding interface between the SARS-CoV-2 RBD and human ACE2 or the SARS-CoV-2 RBD and neutoralizing antibodies.

Hayashi *et al*. S.Figure 4A

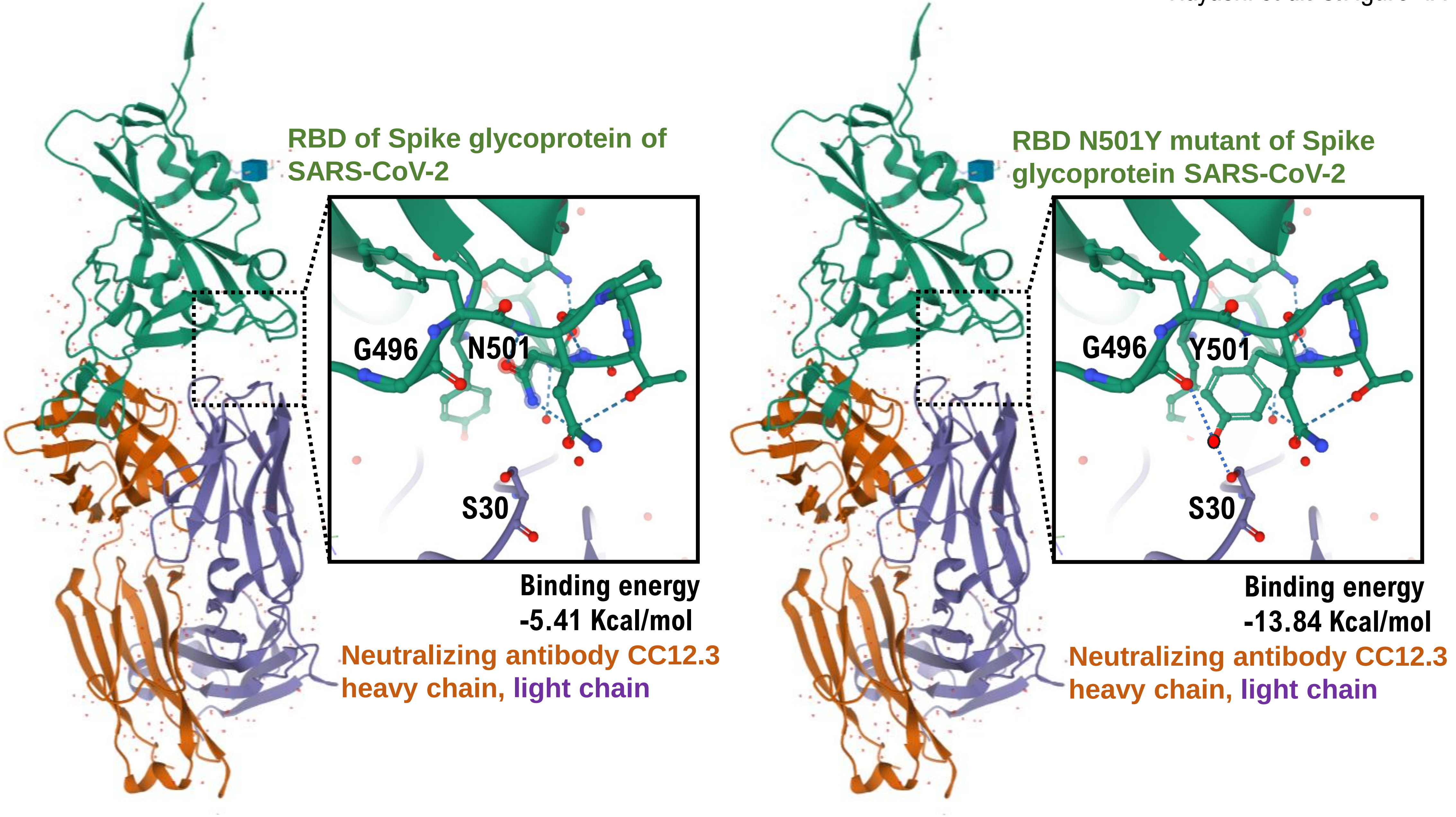

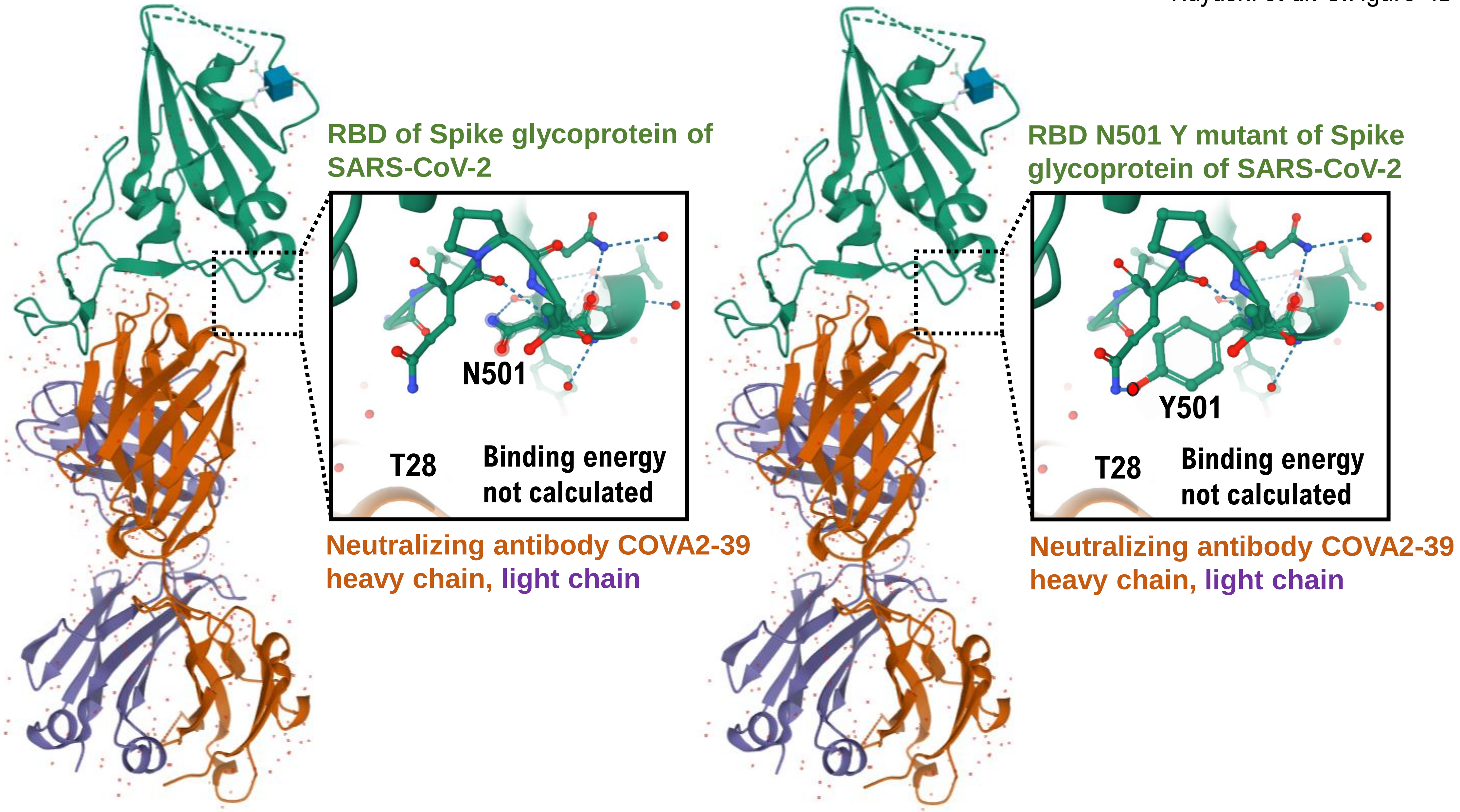
RBD of Spike glycoprotein of SARS-CoV-2
N501
T28
Binding energy not calculated
Neutralizing antibody COVA2-39 heavy chain, light chain
RBD N501 Y mutant of Spike glycoprotein of SARS-CoV-2
Y501
T28
Binding energy not calculated
Neutralizing antibody COVA2-39 heavy chain, light chain

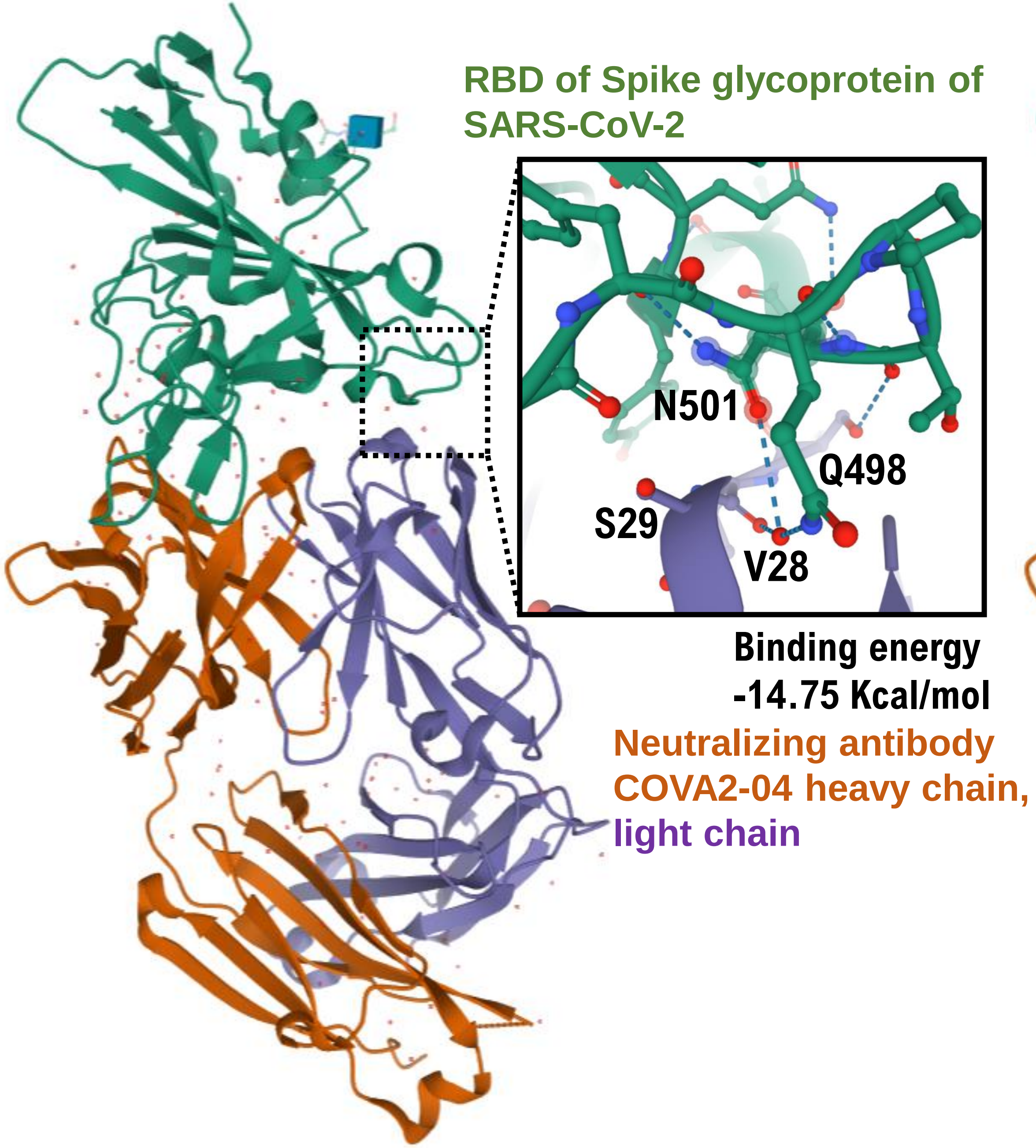
RBD of Spike glycoprotein of SARS-CoV-2
N501
Q498
S29
V28
Binding energy
-14.75 Kcal/mol
Neutralizing antibody COVA2-04 heavy chain,
light chain

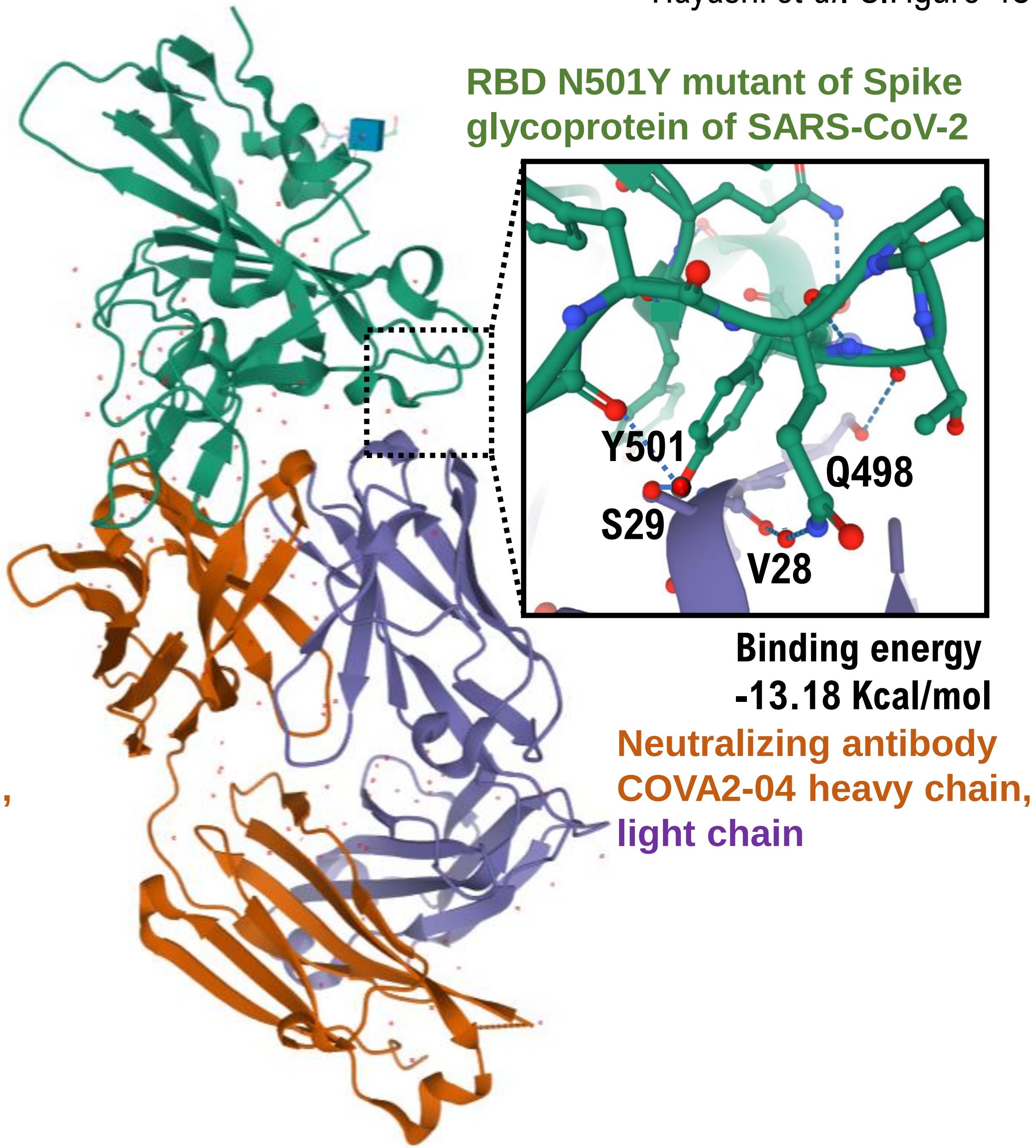
RBD N501Y mutant of Spike glycoprotein of SARS-CoV-2
Y501
Q498
S29
V28
Binding energy
-13.18 Kcal/mol
Neutralizing antibody COVA2-04 heavy chain,
light chain

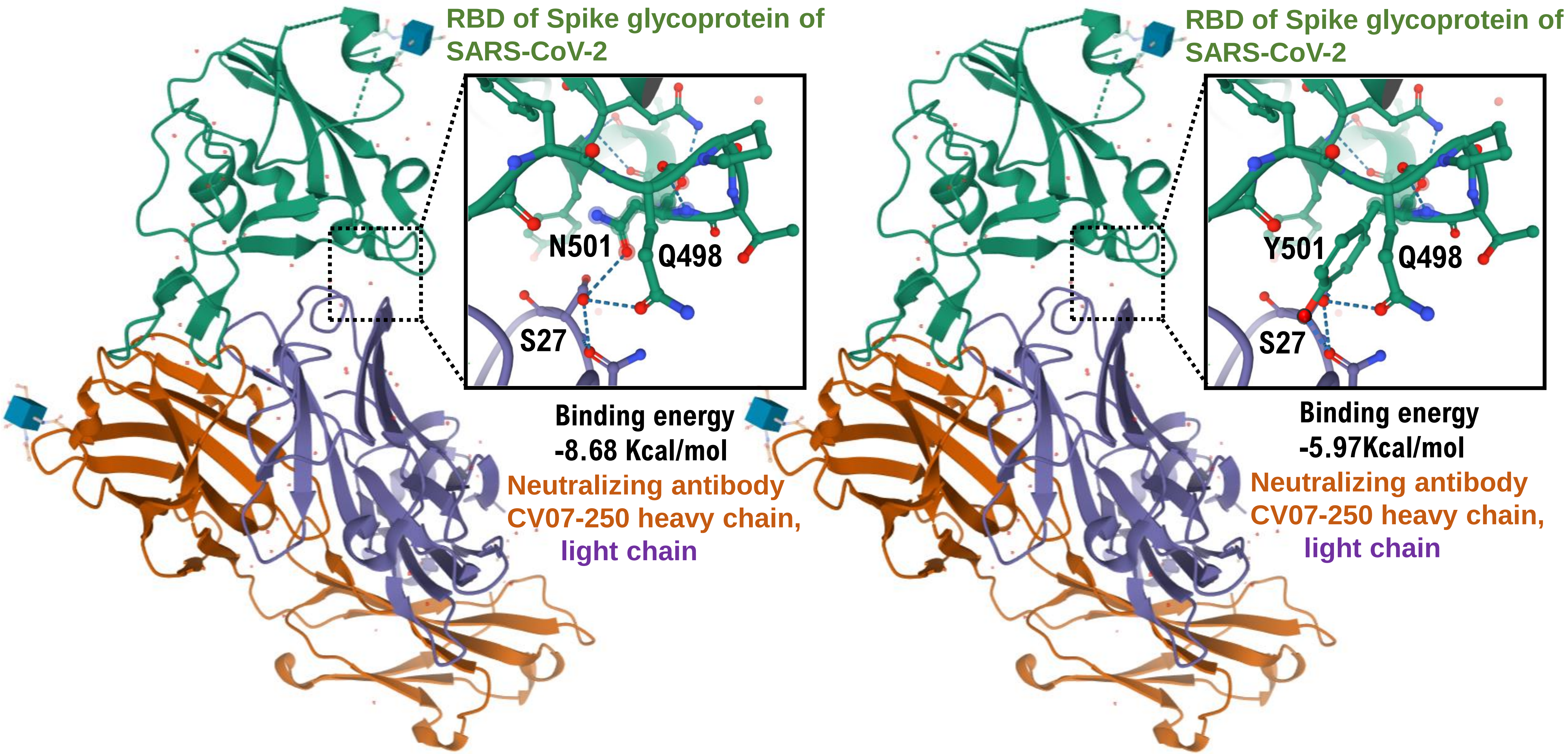
RBD of Spike glycoprotein of SARS-CoV-2
N501
Q498
S27
Binding energy -8.68 Kcal/mol
Neutralizing antibody CV07-250 heavy chain,
light chain
RBD of Spike glycoprotein of SARS-CoV-2
Y501
Q498
S27
Binding energy -5.97Kcal/mol
Neutralizing antibody CV07-250 heavy chain,
light chain

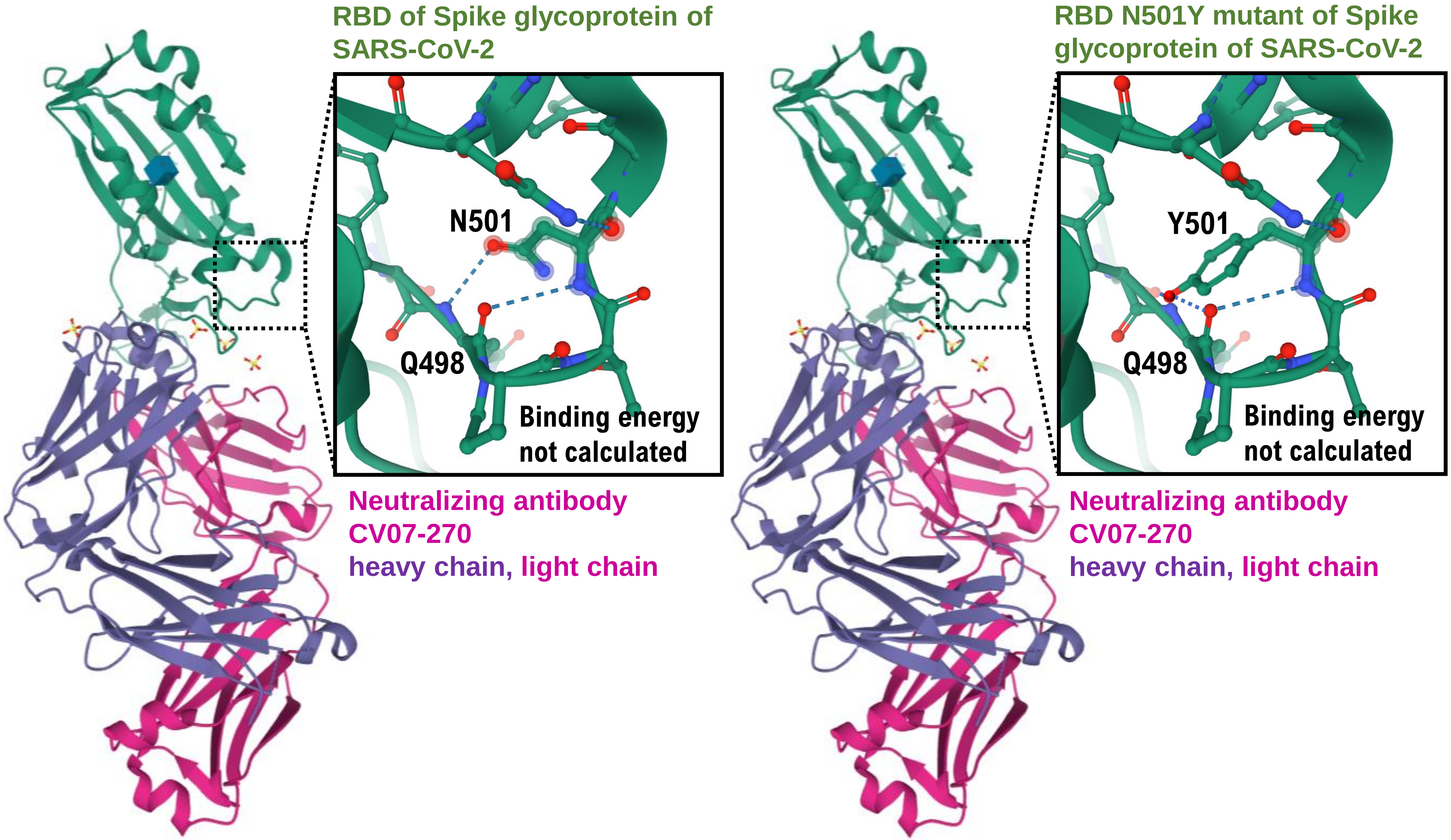
RBD of Spike glycoprotein of SARS-CoV-2
N501
Q498
Binding energy not calculated
Neutralizing antibody CV07-270
heavy chain, light chain
RBD N501Y mutant of Spike glycoprotein of SARS-CoV-2
Y501
Q498
Binding energy not calculated
Neutralizing antibody CV07-270
heavy chain, light chain

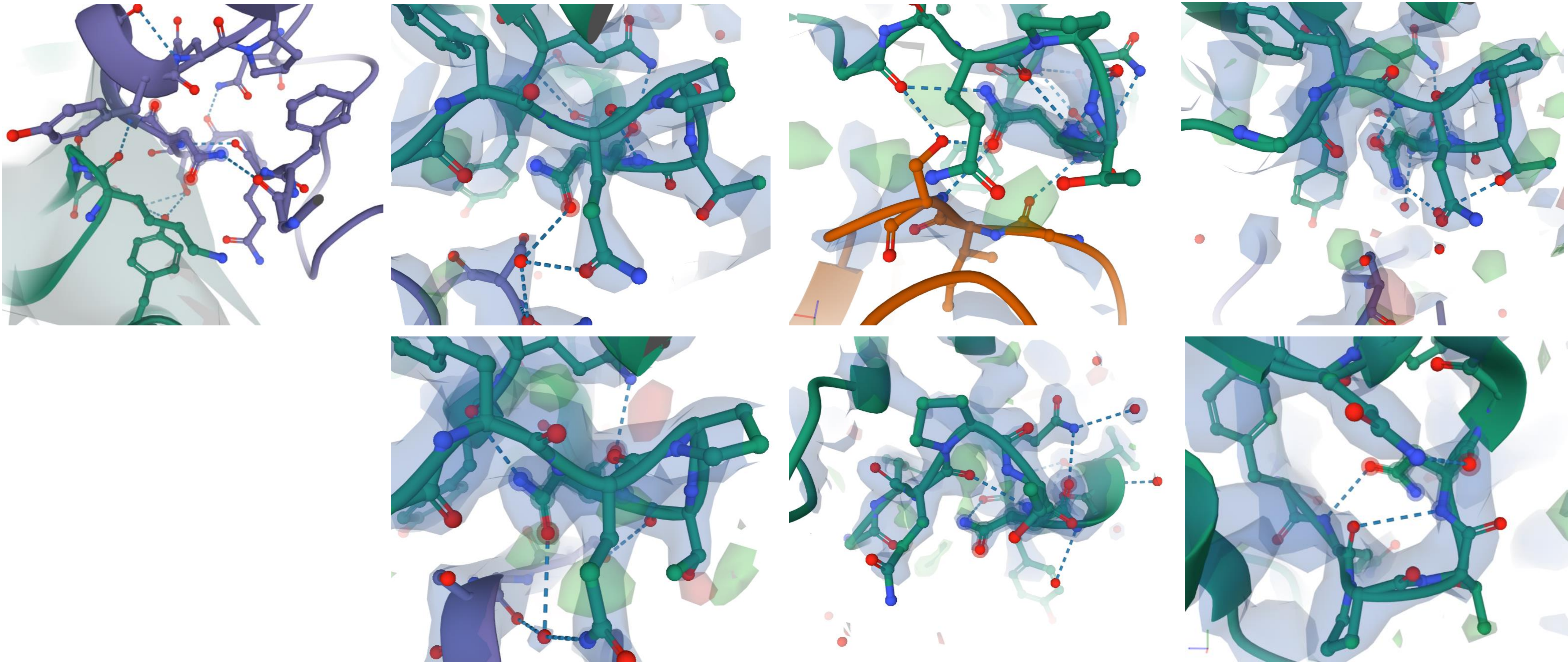

**Electron density map.** 2*F*o−*F*c electron density maps contoured at 1.5σ at the binding interface between the SARS-CoV-2 RBD and human ACE2 or the SARS-CoV-2 RBD and neutoralizing antibodies.

**6XC2 : Crystal structure of SARS-CoV-2 receptor binding domain in complex with neutralizing antibody CC12.1**
Yuan, M., Liu, H., Wu, N.C., Zhu, X., Wilson, I.A. (2020) Science **369**: 1119-1123
**Released** 2020-07-08 **Method** X-RAY DIFFRACTION 3.112 Å
**Organisms** Homo sapiens SARS-CoV-2
**Macromolecule** CC12.1 heavy chain (protein) CC12.1 light chain (protein) Spike protein S1 (protein)

**6XC4** : Crystal structure of SARS-CoV-2 receptor binding domain in complex with neutralizing antibody CC12.3
Yuan, M., Liu, H., Wu, N.C., Zhu, X., Wilson, I.A. (2020) Science **369**: 1119-1123
**Released** 2020-07-08 **Method** X-RAY DIFFRACTION 3.112 Å
**Organisms** Homo sapiens SARS-CoV-2
**Macromolecule** CC12.3 heavy chain (protein) CC12.3 light chain (protein) Spike protein S1 (protein)

**7JMP** : Crystal structure of SARS-CoV-2 receptor binding domain in complex with neutralizing antibody COVA2-39
Wu, N.C., Yuan, M., Liu, H., Zhu, X., Wilson, I.A. (2020) Biorxiv
**Released** 2020-08-26 **Method** X-RAY DIFFRACTION 1.712 Å
**Organisms** Homo sapiens SARS-CoV-2
**Macromolecule** COVA2-39 heavy chain (protein) COVA2-39 light chain (protein) Spike protein S1 (protein)

**7JMO** : Crystal structure of SARS-CoV-2 receptor binding domain in complex with neutralizing antibody COVA2-04
Wu, N.C., Yuan, M., Liu, H., Zhu, X., Wilson, I.A. (2020) Biorxiv
**Released** 2020-08-26 **Method** X-RAY DIFFRACTION 1.712 Å
**Organisms** Homo sapiens SARS-CoV-2
**Macromolecule** COVA2-04 heavy chain (protein) COVA2-04 light chain (protein) Spike protein S1 (protein)

**6XKQ** : Crystal structure of SARS-CoV-2 receptor binding domain in complex with neutralizing antibody CV07-250
Yuan, M., Liu, H., Zhu, X., Wu, N.C., Wilson, I.A. (2020) Cell
**Released** 2020-08-26 **Method** X-RAY DIFFRACTION 2.55 Å
**Organisms** Homo sapiens SARS-CoV-2
**Macromolecule** CV07-250 heavy chain (protein) CV07-250 light chain (protein) Spike protein S1 (protein)

**6XKP** : Crystal structure of SARS-CoV-2 receptor binding domain in complex with neutralizing antibody CV07-270
Liu, H., Yuan, M., Zhu, X., Wu, N.C., Wilson, I.A. (2020) Cell
**Released** 2020-08-26 **Method** X-RAY DIFFRACTION 2.72 Å
**Organisms** Homo sapiens SARS-CoV-2
**Macromolecule** CV07-270 heavy chain (protein) CV07-270 light chain (protein) Spike protein S1 (protein)

**6XGD:** Complex of SARS-CoV-2 receptor binding domain with the Fab fragments of two neutralizing antibodies **REGN-COV2**
**Released:** 2020-06-24 **Method:** ELECTRON MICROSCOPY **Resolution:** 3.90 Å
**Organism(s):** Severe acute respiratory syndrome coronavirus 2, Homo sapiens
**DOI:** 10.2210/pdb6XDG/pdb **EMDataResource:** EMD-22137
